# Intraoperative arterial blood pressure waveform morphology variability is associated with acute kidney injury after cardiac surgery

**DOI:** 10.1101/2025.10.12.25337819

**Authors:** Cheng-Hsi Chang, Shen-Chih Wang, Wen-Kuei Chang, Chien-Kun Ting, Ming-Jen Lu, Chia-Hsun Lin, Chai-Hock Chua, Hau-Tieng Wu, Yu-Ting Lin

## Abstract

**Purpose:** Postoperative acute kidney injury (post-AKI) remains common after cardiac surgery with cardiopulmonary bypass (CPB). Conventional blood pressure (BP)-related measures may overlook hemodynamic information contained in the arterial blood pressure waveform (ABP). We hypothesize that intraoperative ABP morphology variability, quantified by VarM, captures the hemodynamic impact of CPB beyond its duration and may improve intraoperative post-AKI prediction.

**Methods:** We conducted a prospective observational cohort study at a tertiary medical center enrolling adults undergoing elective cardiac surgery with CPB between November 2019 and November 2022. VarM was calculated from the intraoperative ABP using an unsupervised manifold-learning algorithm. Post-AKI occurring within two weeks was defined according to Kidney Disease: Improving Global Outcomes (KDIGO) serum creatinine criteria. Logistic regression evaluated VarM’s predictive value, its complementarity to CPB duration, and the performance of a combined model for post-AKI prediction.

**Results:** Of 101 patients enrolled, 18 were excluded for preoperative end-stage kidney disease requiring dialysis. Subjects developing post-AKI had lower VarM and slower recovery after CPB. Post-CPB VarM was inversely associated with postoperative creatinine (Spearman correlation coefficient, -0.387[-0.597,-0.152], P=0.0029). VarM achieved an area-under-curve (AUC) of 0.747 for post-AKI prediction, outperforming eGFR (0.624) and EuroSCOPE (0.623), and similar to CPB duration (0.756). A combined VarM-CPB duration model achieved an AUC of 0.817, significantly outperforming either variable alone (P=0.0025).

**Conclusion:** Post-CPB VarM was associated with post-AKI, and provided complementary information to CPB duration. Combining both measures may improve intraoperative post-AKI prediction.

## Introduction

Acute kidney injury (AKI) is a common complication after cardiac surgery and is associated with significant morbidity and mortality^1-4^. Several preoperative and intraoperative factors have been associated with postoperative AKI (post-AKI), including baseline estimated glomerular filtration rate (eGFR) ^5^, operative risk assessed by the European System for Cardiac Operative Risk Evaluation (EuroSCORE) ^6^, aortic cross-clamp and cardiopulmonary bypass (CPB) duration^7^, oxygen delivery and perfusion parameters^8^, hemolysis^9^, and intraoperative blood pressure (BP) characteristics^10-14^. Among these factors, this study focuses on intraoperative BP and hypotension burden. It has been known that hypotension burden is consistently associated with post-AKI risk^15^, while strategies targeting avoidance of intraoperative hypotension have not consistently reduced post-AKI incidence^16,17^. These findings suggest that conventional BP-derived measures may not fully capture the hemodynamic information relevant to postoperative renal injury.

Conventional BP-related measures, including systolic BP (SBP), diastolic BP (DBP), pulse pressure (PP), and BP variability (BPV), are widely used in clinical practice, but capture only selected features of the arterial blood pressure waveform (ABP). These measures are, in essence, low-dimensional summaries of a complex physiologic signal. Increasingly, investigators have sought to characterize hemodynamic properties encoded in ABP morphology^18-21^. Notably, although morphological differences between consecutive ABP pulses may be subtle, their cumulative temporal patterns can encode complex cardiovascular information^19,22^. Emerging evidence suggests that greater variability in ABP morphology is associated with more favorable physiologic and clinical states^18,23-25^. Such variability may reflect the integrated effects of multiple cardiovascular regulatory mechanisms, analogous to the information captured by heart rate variability (HRV)^26^ and breathing pattern variability.^15^

Whether intraoperative ABP morphology variability is associated with post-AKI after cardiac surgery remains unknown^27^. We hypothesized that greater variability in ABP morphology over the post-CPB period, quantified by the index VarM, is associated with lower post-AKI. This association is via encoded hemodynamic impacts of CPB that are complementary to CPB duration. We apply logistic regression to evaluate VarM’s predictive value beyond established preoperative and intraoperative risk factors and its complementarity to CPB duration. We then consider a combined model incorporating post-CPB VarM and CPB duration for post-AKI prediction.

## Methods

### Patients

This single-center prospective observational cohort study was conducted at Shin Kong Wu Ho-Su Memorial Hospital after approval by the institutional review board (IRB) (20181205R) and in accordance with the Declaration of Helsinki. Data analysis was written and filed with an IRB before data were accessed; while statistical plan of AKI association was written after the data were accessed.

Adult patients undergoing elective cardiac surgery with CPB, including coronary artery bypass graft (CABG), valvular surgery, or combined CABG and valvular surgery, were enrolled after providing written informed consent between November 2019 and November 2022. Surgeries were performed by three cardiac surgeons (MJL, CHL, CHC). Patients were excluded if they required renal replacement therapy, had a pacemaker, an intra-aortic balloon pump, or extracorporeal membrane oxygenation, or if they had significant cardiac arrhythmias, such as atrial fibrillation or frequent premature ventricular contractions.

Anesthesia was maintained with desflurane, and mean arterial pressure (MAP) during CPB was maintained between 50 and 70 mmHg. Data acquisition was fully automated and did not interfere with routine anesthetic management. After surgery, patients were routinely transferred to the intensive care unit (ICU), where complete blood counts and serum chemistry panels were obtained daily according to routine clinical practice and at the discretion of the treating physician. Serum creatinine was measured preoperatively and postoperatively on the day of surgery (postoperative day (POD) 0) and on POD1-POD3 as part of routine institutional care. Invasive hemodynamic monitoring included ABP via a radial arterial catheter and pulmonary artery catheterization or central venous catheterization, according to the clinical indication. Aminoglycosides were not used as prophylactic antibiotics. The preoperative EuroSCORE was recorded as a measure of baseline operative risk.

### Data collection

Continuous physiological waveform as well as all numeric data were collected from the patient monitor (IntelliVue MX800, Philips, Amsterdam, Netherlands) using the data collection software ixTrend (ixellence GmbH, Wildau, Germany). ABP waveforms were obtained from the direct radial arterial cannulation uniformly sampled at 125 Hz. The anesthesiologist (CHC) logged the anesthetic events and surgical progress with an automated electronic system.

We defined seven surgical periods: P1, before anesthetic induction; P2, a 10-minute period during central venous catheter insertion; P3, a 10-minute period during vascular graft harvest; P4, 0-10 minutes after weaning from CPB; P5, 30-40 minutes after weaning from CPB; P6, from the end of CPB to 10 minutes after sternal closure; and P7, 10 minutes after completion of sternal closure. We analyzed several time periods after weaning from CPB due to the significant impact of CPB on AKI^7^.

### Renal function association and primary outcome

Preoperative eGFR was used as a measure of baseline renal function. The primary outcome was post-AKI occurring within two weeks after surgery and was treated as a binary outcome (post-AKI present or absent). AKI was defined according to the modified KDIGO criteria^28^, as an increase in serum creatinine to ≥1.5 times the baseline value or an absolute increase of ≥0.3 mg/dl. Patients with end-stage renal disease (ESRD) requiring renal replacement therapy before surgery were excluded from the AKI analysis.

### Quantifying variability of morphology

ABP morphology variability was quantified using DDmap^18^, an unsupervised manifold-learning technique^29,30^. Individual pulse cycles were automatically identified using the point of maximum upstroke velocity during the ascending phase of each ABP pulse as the fiducial point^19^. For each cardiac cycle, the preceding nadir was defined as the waveform origin, and the resulting pulse waveform was represented as a high-dimensional vector.^19^ (Fig. 1). To minimize the influence of absolute BP levels, which were continuously monitored and managed during cardiac surgery, the DBP was subtracted from each pulse waveform. DDmap was then applied to the resulting normalized pulse waveforms to eliminate the respiratory pulse pressure variation and derive VarM^18^. VarM quantifies how the consecutive pulse waveforms change within the learned low-dimensional manifold, thereby capturing cumulative variability in waveform morphology. It is derived solely from the ABP waveform and does not require additional clinical variables as inputs. The DDmap-based approach is robust to subtle changes in waveform morphology and provides a compact representation of their cumulative variability over time.^22^ VarM was normalized to a 0-100 scale^31^. Define the slope of VarM to be the VarM between surgical periods P2 and P6.

**Figure 1.**
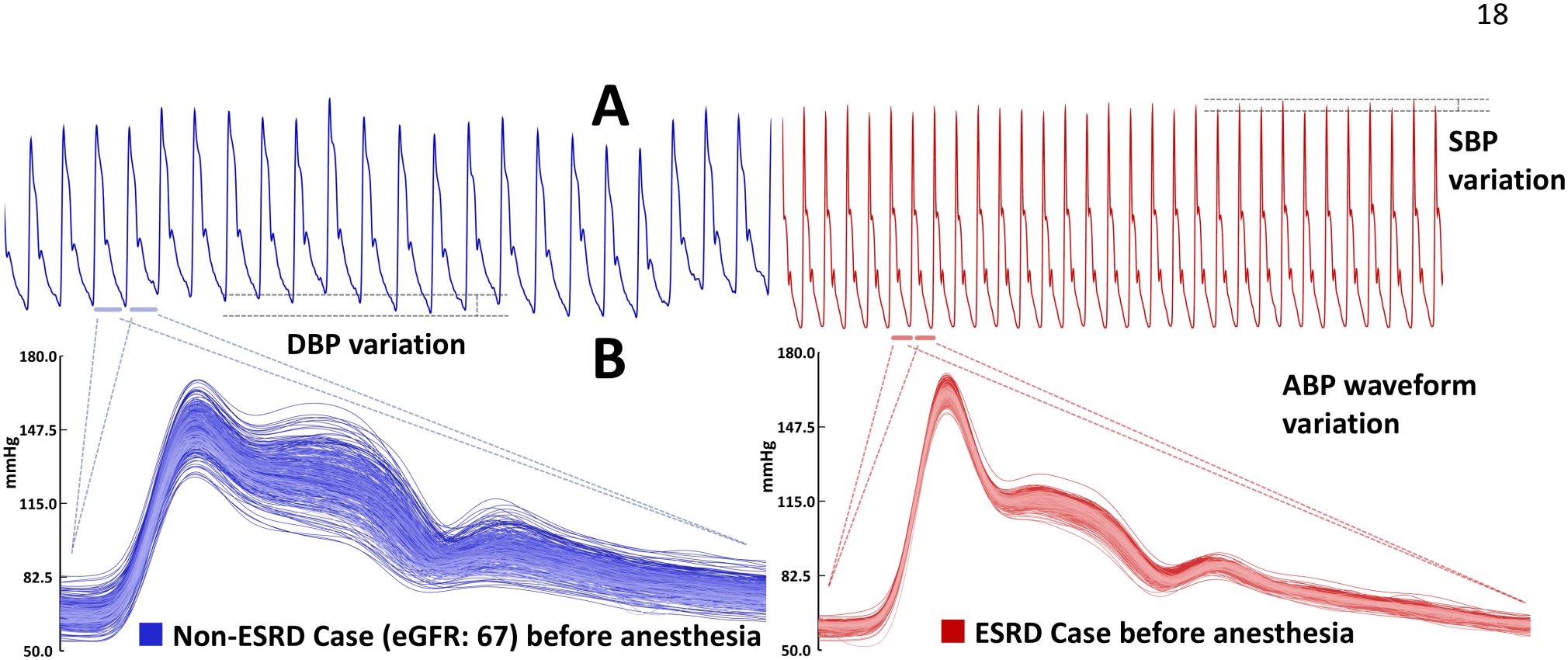
ABP waveform data from two cases before anesthetic induction demonstrate different variations in morphology, with better renal function associated with greater variation. Although this difference may not be readily apparent in the raw waveform tracing (panel A), subtle yet intricate morphologic changes are evident when examined on a per-pulse basis (panel B). From the perspective of waveform morphology, variations in DBP and SBP reflect only partial aspects of VarM. ABP, arterial blood pressure; DBP, diastolic blood pressure; ESRD, end-stage renal disease; SBP, systolic blood pressure; VarM, variation of ABP waveform morphology.

### Basic vital parameters and the variation indices

Conventional BP measures, including SBP, DBP, MAP, and PP, were derived from each pulse cycle. From the resulting beat-to-beat measurements, we calculated the mean and established indices of BP variability (BPV)^14,32,33^, including standard deviation (SD), coefficient of variation (CV), average real variability (ARV), interquartile range (IQR), and quartile coefficient of dispersion (QCD) of SBP, DBP, MAP, and PP. Pulse rate, a surrogate of heart rate, was also derived from ABP, with corresponding variability indices calculated as surrogates of HRV.

Hypotension burden was quantified as the time-weighted average of MAP below 65 mmHg (TWA-MAP <65 mmHg)^34^. After artifact removal, TWA-MAP <65 mmHg was calculated as the area between the 65-mmHg threshold and the MAP curve during the period when MAP was below 65 mmHg, divided by the total duration of the analyzed period. These measures were used solely as comparator variables in association and prediction analyses of post-AKI.

### Statistical analysis

Continuous demographic and laboratory variables were reported as median (interquartile range [IQR]). Missing data were handled using pairwise deletion for univariate analyses and complete-case deletion for multivariable analyses.

Spearman correlation coefficient (SCC) was used to explore association. Confidence intervals (CIs) for correlation analysis were estimated using 100,000 bootstrap resampling. The Wilcoxon rank-sum exact test was used for between-group comparisons. Statistical significance was defined as P<0.05, with Bonferroni correction applied for multiple comparisons.

Univariate logistic regression models were used to evaluate the predictive value of VarM, eGFR, EuroSCORE, and CPB duration for post-AKI. To evaluate whether VarM provided complementary information beyond CPB duration, we compared a bivariable logistic regression model incorporating both predictors with the corresponding univariate models. Model differences were evaluated using likelihood-ratio tests for nested models. Predictive performance was evaluated using leave-one-out and four-fold cross-validation, with the synthetic minority oversampling technique (SMOTE) used to address class imbalance^35^. Area under the curve (AUC) and 95% CIs estimated from 100,000 bootstrap resamples were reported. Accuracy and precision were calculated at the threshold maximizing the Youden index.

All the statistical analyses were carried out with R (version 4.5.0; the R Foundation for Statistical Computing).

## Results

### Epidemiology

From 116 enrolled patients undergoing CABG and/or valvular surgery requiring CPB support, we excluded 15 cases due to suboptimal data quality. We analyzed baseline data from 101 patients (83 males and 18 females; Age 64.6 [59,70.0]) and evaluated AKI outcome from 83 non-ESRD cases (Fig. 2). There were 27 post-AKI cases (32.5%) who presented higher creatinine postoperatively (POD1: 1.55 [1.31,2.08], POD2: 1.19 [0.9,1.4], POD3: 1.2 [0.92,1.42], unit: mg/dL) than 56 non-post-AKI cases (POD1: 0.94 [0.84,1.17], POD2: 0.75 [0.63,0.86], POD3: 0.75 [0.66,0.89]).

**Figure 2.**
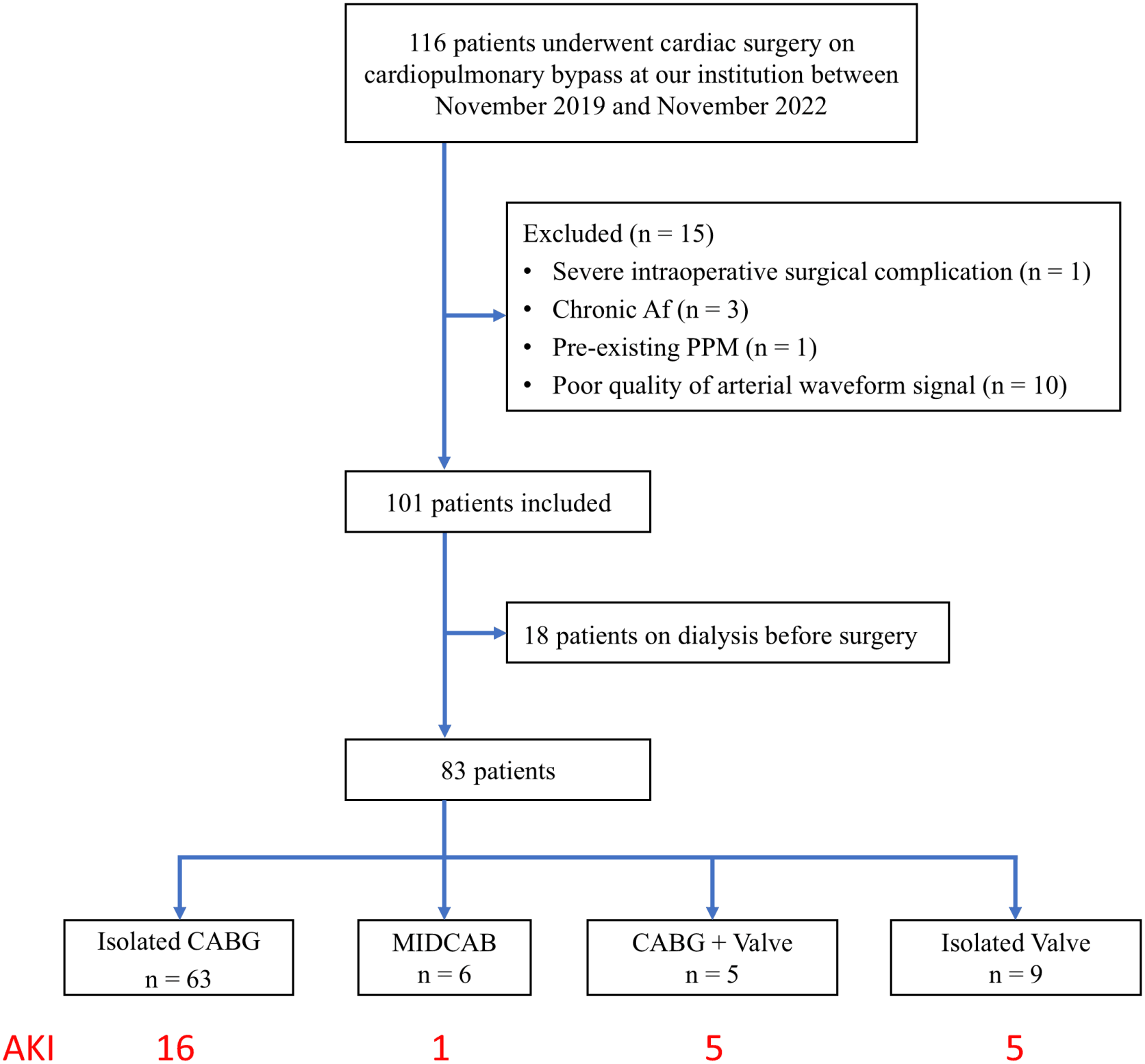
Flowchart of patient selection. AKI incidence was higher when valvular procedure was involved. Af: atrial fibrillation. CABG: coronary artery bypass graft. MIDCAB: minimally invasive direct coronary artery bypass. PPM: permanent pacemaker.

### Association of pre-operative factors with postoperative AKI

Post-AKI was not associated with the baseline eGFR (SCC=-0.185 [-0.387,0.0346], p=0.094), neither the EuroSCORE (SCC=0.191[-0.041,0.400], p=0.088).

### Association of intraoperative factors with postoperative AKI

Intraoperative factors associated with post-AKI include longer CPB time (SCC=0.415 [0.20, 0.58], p=0.0001) and valvular surgery (SCC=0.41 [0.17, 0.60], p=0.0001). The vasopressor dosage rates at the end of surgery showed marginal associations for Norepinephrine (SCC=0.26 [0.036,0.477], p=0.016) and Dopamine (SCC=0.27 [0.053,0.47], p=0.012). The hypotension measured as TWA-MAP <65 mmHg showed no association with AKI.

Other physiological parameters in the corresponding surgical periods, including various BP measures, heart rate, and their derived variations, were not associated with either baseline renal function or post-AKI.

### Association of VarM with renal function

VarM was highest during P1 and decreased afterward in both post-AKI and non-post-AKI groups until CPB (Fig. 3A). VarM during P1 presented no association with baseline eGFR (SCC=0.196 [-0.005,0.386], p=0.062) and post-AKI (SCC=0.10 [-0.123,0.313], p=0.39).

**Figure 3.**
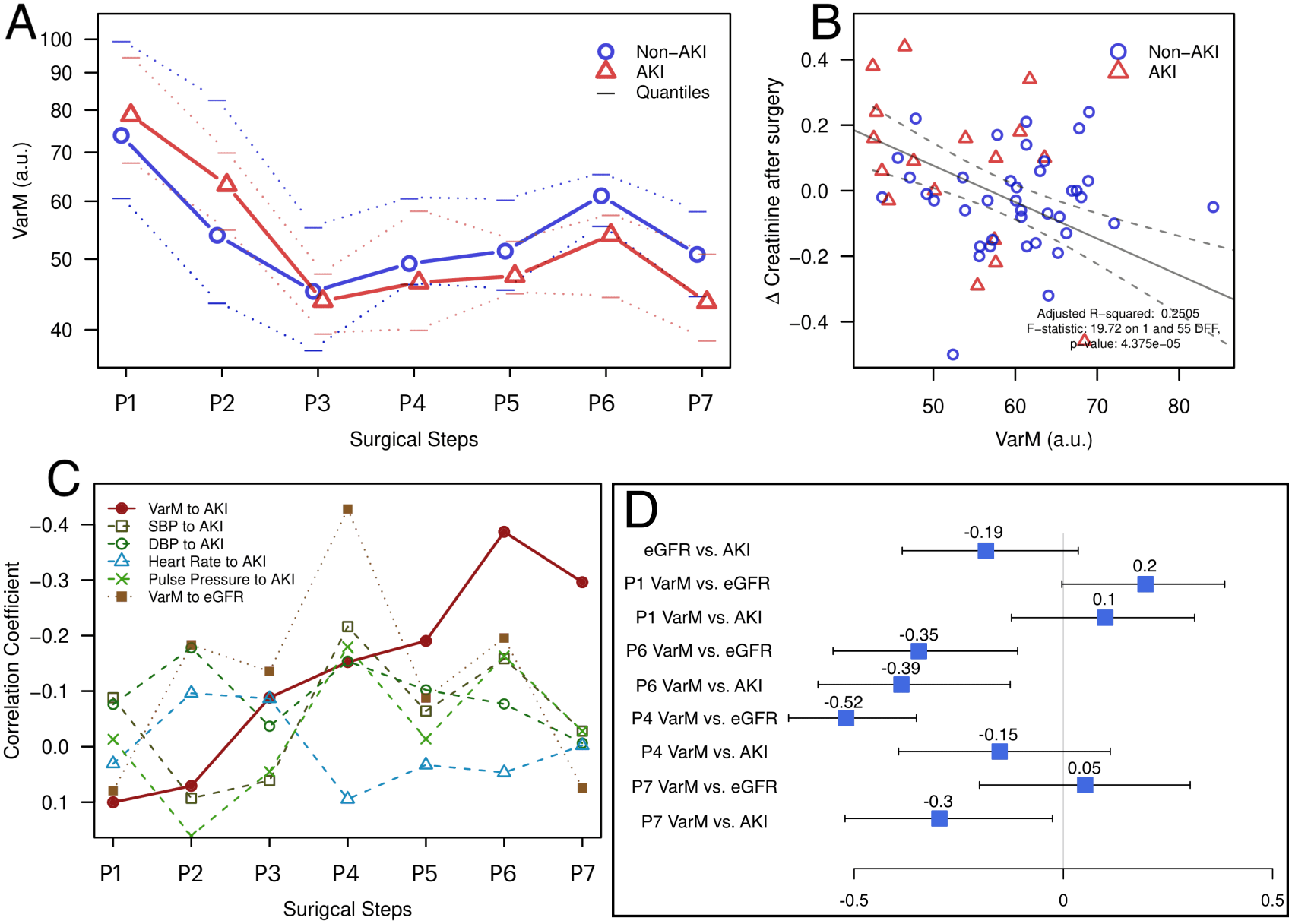
Panel A: Dynamics of VarM across surgery phases. After weaning from CPB, VarM remained lower in patients who developed AKI than in those who did not (panel A). Panel B: The association between VarM and the change in serum creatinine on POD1, with linear regression lines and 95% confidence intervals superimposed on the scatterplots. Panel C: SCC of VarM and conventional hemodynamic measures with eGRF and AKI. Panel D: SCC of VarM over different surgical periods, particularly P1, P4, P6, and P7, with eGFR and AKI and their corresponding 95% confidence intervals. ABP: arterial blood pressure; AKI: acute kidney injury; AUC: area under the receiver operating characteristic (ROC) curve; CPB: cardiopulmonary bypass; CV Cath: central venous catheter; DBP: diastolic blood pressure; eGFR: estimated glomerular filtration rate; SBP: systolic blood pressure; SCC: Spearman correlation coefficient; VarM: variability of ABP waveform morphology.

The postoperative creatinine increase was negatively associated with VarM following CPB weaning (Fig. 3B), with low SCC values observed during P1-P4.

After weaning from CPB, VarM increased toward sternal closure (VarM slope=72.24, p=0.0006), with higher VarM in the non-post-AKI group than in the post-AKI group (p=0.003; Fig. 3A). Lower VarM slope was also associated with post-AKI (p=0.0001).

The association between VarM and post-AKI across surgical periods surrounding CPB are presented in Fig. 3D. VarM was negatively associated with baseline eGFR during P4 (SCC=-0.519 [-0.656,-0.349], p=5.77 × 10^−7^) and P6 (SCC=-0.345

[-0.552,-0.108], p=0.004), but not during P7. During P6, VarM showed its strongest association with post-AKI (SCC=-0.39 [-0.597,-0.152], p=0.0029), compared with other vital parameters examined. This association was weaker but remained statistically significant during P7 (SCC=-0.30 [-0.52,-0.021], p=0.023).

### Postoperative AKI prediction

Univariate logistic regression models based on preoperative indices yields AUCs of 0.624 [0.519,0.728] for eGFR and 0.626 [0.521,0.731] for EuroSCORE (Fig.5A and Fig.5B).

**Figure 4.**
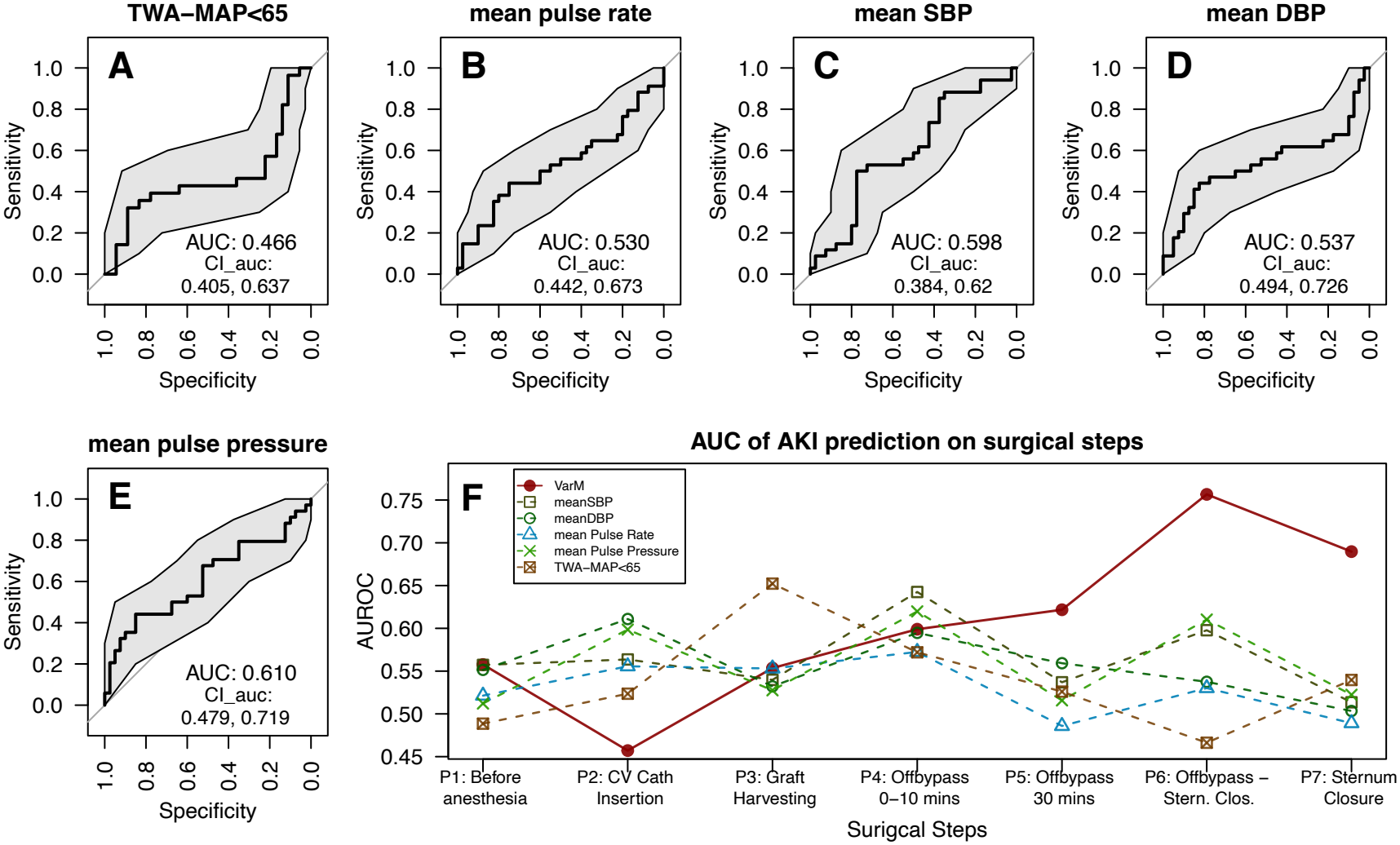
Panels A to E: ROCs of univariate prediction of post-AKI using various indices calculated from P6. From A to E, the predictors are TWA-MAP<65 mmHg, pulse rate, mean SBP, mean DBP, mean PP, respectively. Panel F: AUC of univariate prediction of post-AKI using various indices calculated from different surgical periods. AKI: acute kidney injury; AUC: area under the curve; CPB: cardiopulmonary bypass; DBP: diastolic blood pressure; PP: pulse pressure; SBP: systolic blood pressure; VarM: variability of ABP waveform morphology.

**Figure 5.**
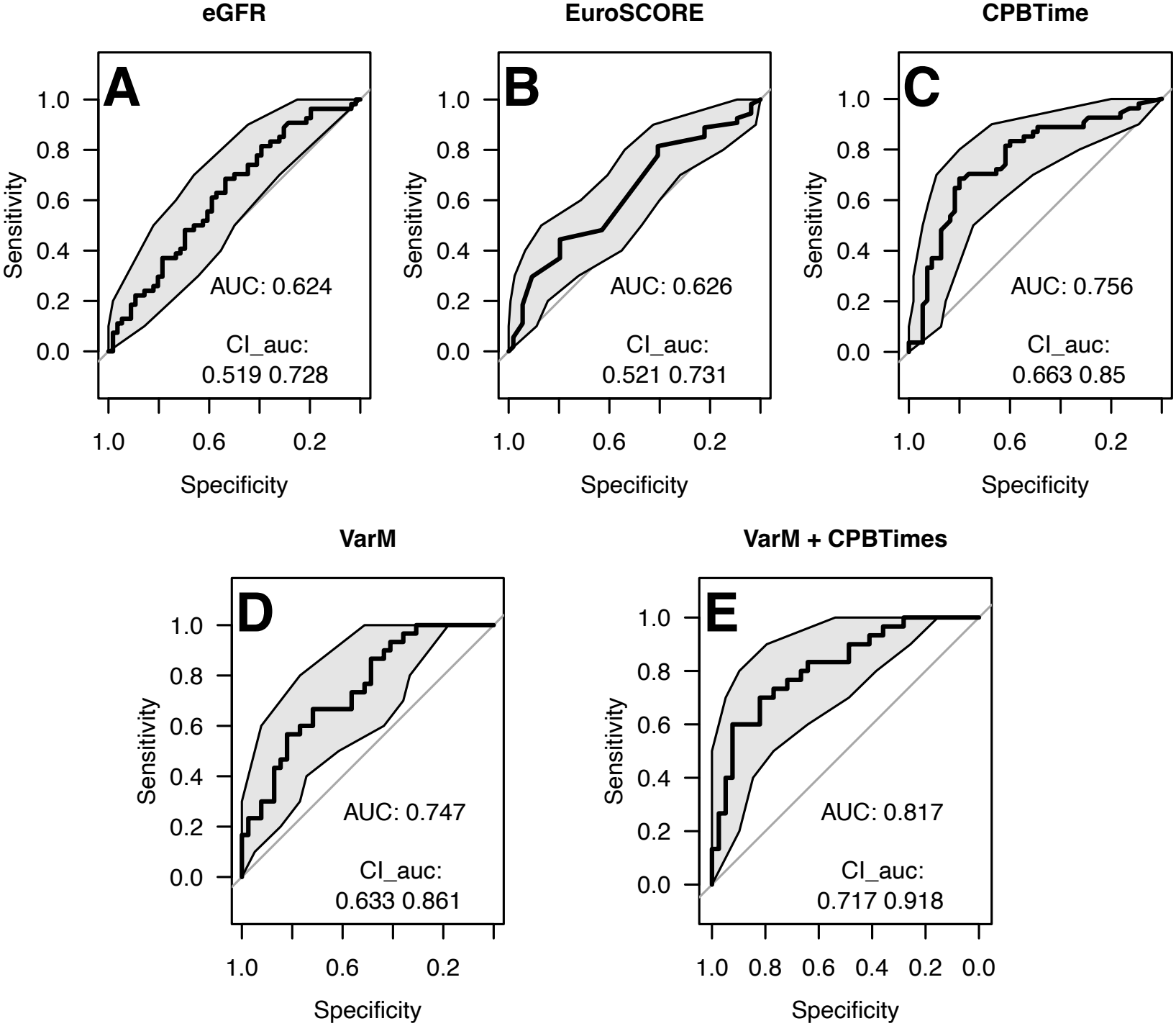
ROC of logistic regression analyses with their 95% confidence interval and AUC. From A to E are eGFR, EuroSCORE, CPB time, post-CPB VarM, and the combination of CPB time and post-CPB VarM. ABP: arterial blood pressure; AKI: acute kidney injury; AUC: area under the curve; CPB: cardiopulmonary bypass; eGFR: estimated glomerular filtration rate; VarM: variability of ABP waveform morphology.

Among conventional hemodynamic indices, PP had the highest AUC of 0.610 [0.479,0.719], whereas TWA-MAP<65 mmHg showed limited discrimination of AUC 0.466 [0.405,0.637] (Fig.4E and Fig.4A). Among HRV and BPV indices (see Figure S1 in Supplementary Material), ARV of DBP achieves the best performance with AUC 0.695 [0.573,0.817].

Regarding VarM, it consistently outperformed conventional measures in discriminating post-AKI across P5-P7 (Fig.4G), with the highest discriminative performance observed during P6 of AUC 0.747 [0.633,0.861] (Fig.5D). On the other hand, CPB duration yielded AUC of 0.756 [0.663,0.85] (Fig.5C). A bivariable logistic regression model incorporating both VarM during P6 and CPB duration achieved an AUC of 0.817 [0.717,0.918] (Fig.5E), significantly outperforming the individual univariate models (p=0.0025). These findings suggest that VarM during P6 provides predictive information for post-AKI complementary to CPB duration.

## Discussion

In this prospective observational study, intraoperative ABP morphology variability, quantified by VarM, was associated with post-AKI after cardiac surgery with CPB. VarM was highest before anesthetic induction and progressively decreased during surgery, with its association with AKI becoming more pronounced after CPB weaning. Patients who developed AKI had lower VarM after CPB weaning and a less pronounced post-CPB increase. Importantly, combining VarM after CPB weaning and CPB duration significantly improved intraoperatively post-AKI prediction.

Our findings suggest that intraoperative VarM provides post-AKI information beyond conventional hemodynamic measures. CPB imposes substantial physiological stress through alternations in blood flow, oxygen delivery, and systematic inflammatory responses, while prolonged CPB duration has been associated with an increased risk of post-AKI.^7^ Although CPB duration reflects the length of exposure to these stressors, it does not fully capture individual differences in the physiological response to CPB or subsequent recovery. We hypothesize that VarM, by quantifying beat-to-beat variability in ABP morphology, captures aspects of this integrated physiological response. The observed increase in VarM after weaning from CPB, together with lower VarM in patients who developed post-AKI, may indicate differences in the recovery of physiological regulatory dynamics. This interpretation is consistent with the broader concept that patterns of physiological variability^36^ can provide insight into the function of complex biological systems and their responses to stress. A similar pattern was observed in our previous study of liver transplantation, in which higher VarM during the neohepatic phase was associated with better clinical outcomes^18^. Taken together, these findings suggest that ABP morphology variability may reflect aspects of systemic physiological resilience across different organs and surgical settings, warranting further investigation.

The AKI incidence (32.5%) in this study was comparable to that reported in previous studies (25-45%)^37^. The pathophysiology of post-AKI after cardiac surgery involves inflammation, hypoperfusion, and CPB-related stress^38-40^, processes closely linked to cardiovascular function. Extreme BP values may reflect adverse cardiovascular states: hypotension may indicate inadequate organ perfusion^11,40-43^, whereas hypertension may reflect arterial stiffness or other adverse vascular conditions^12^. However, conventional BP measures and derived indices, including BPV and hypotension burden, showed lower predictive performance for post-AKI than VarM (Supplementary Table S1). Mitrev et al. reported that pre-CPB PP predicted post-AKI, whereas PP during the post-CPB period and the first ICU hour did not^21^. McIlroy et al.^44^ reported that they cannot establish association between increasing time spent in any MAP range band during the first 24 hours postoperatively and AKI. Srivastava et al.^45^ showed that greater postoperative decreases in MAP from preoperative baseline was associated with post-AKI. Other commonly reported intraoperative factors, including vasopressor use, surgical type, and CPB duration^37,46-49^, likewise did not provide comparable predictive performance. It is worth noting that TWA-MAP<65 mmHg showed no association with post-AKI. We speculated that the clinical BP management shifted the statistical associations to the dosages of vasopressor medication. Collectively, these findings suggest that VarM captures information relevant to post-AKI risk that is not fully reflected by conventional hemodynamic measures or other commonly used intraoperative variables.

The improvement in post-AKI prediction achieved by combining post-CPB VarM and CPB duration suggests that VarM provides post-AKI information complementary to CPB duration. Together, these variables may enable AKI risk assessment before completion of surgery. Because VarM is derived from routinely recorded ABP waveforms without requiring additional clinical measurements, it may be readily integrated into perioperative monitoring and potentially help anesthesiologists and surgeons for clinical decision-making.

From a signal-processing perspective, VarM and conventional BP measures, including SBP, DBP, MAP, and their variability, are all summary measures of ABP dynamics, but they capture fundamentally different aspects of the signal. Conventional BP measures count on point information of continuous ABP waveform, while VarM characterizes variability in the morphology of the entire pulse waveform. Physiologically, ABP morphology reflects the integrated effects of ventricular ejection, arterial stiffness and compliance, vascular geometry, wave propagation and reflection, and peripheral vascular properties. These features emerge from the continual interaction of multiple cardiovascular regulatory mechanisms that maintain hemodynamic homeostasis^19^. A useful physical analogy is to view the heartbeat as striking the cardiovascular system like a stick striking a drum: the resulting waveform is shaped by the properties of the system that produces and transmits the pulse. In this sense, waveform morphology provides a functional signature of the cardiovascular system, motivating the question of whether one can “hear the shape of a drum.” This concept underlies the development of VarM. Growing evidence supports its clinical relevance, with VarM associated with outcomes after liver transplantation and survival in septic shock^18,24^. Moreover, photoplethysmography, which captures a related peripheral pulsatile signal, may provide similarly informative waveform-level information^25^.

Several limitations should be acknowledged. First, this was the first prospective evaluation of VarM in relation to post-AKI after cardiac surgery. External validation in larger cohorts is needed. Since continuous ABP waveform data are not routinely recorded, retrospective validation may be challenging, and large-scale prospective studies are therefore warranted. Second, the pathophysiological mechanisms linking VarM to post-AKI remain unclear and are likely to involve multiple interacting processes. Future studies incorporating blood and urinary biomarkers may help elucidate these mechanisms and clarify how VarM relates to other clinical variables. Third, all surgeries were performed by three experienced cardiac surgeons. Although this provided procedural consistency, studies involving different institutions, surgical teams, and geographic settings are needed to establish generalizability. Fourth, our standardized anesthetic management provided controlled conditions for VarM measurement, which may limit its application outside the operating room. In routine clinical settings, waveform artifacts related to catheter or tubing kinking, damping, and patient motion may affect VarM estimation and will require appropriate signal-quality assessment and artifact handling.

## Conclusion

Morphology variability in the ABP morphology after weaning from CPB is associated with post-AKI in cardiac surgery. Lower VarM with a decreased trend between weaning from CPB and sternal closure is associated with post-AKI. Combining VarM and CPB duration significantly improved the prediction of post-AKI.

## Data Availability

All data produced in the present study and the R-script for statistical analysis are available upon reasonable request to the authors

---

**Supplementary Figure S1.**
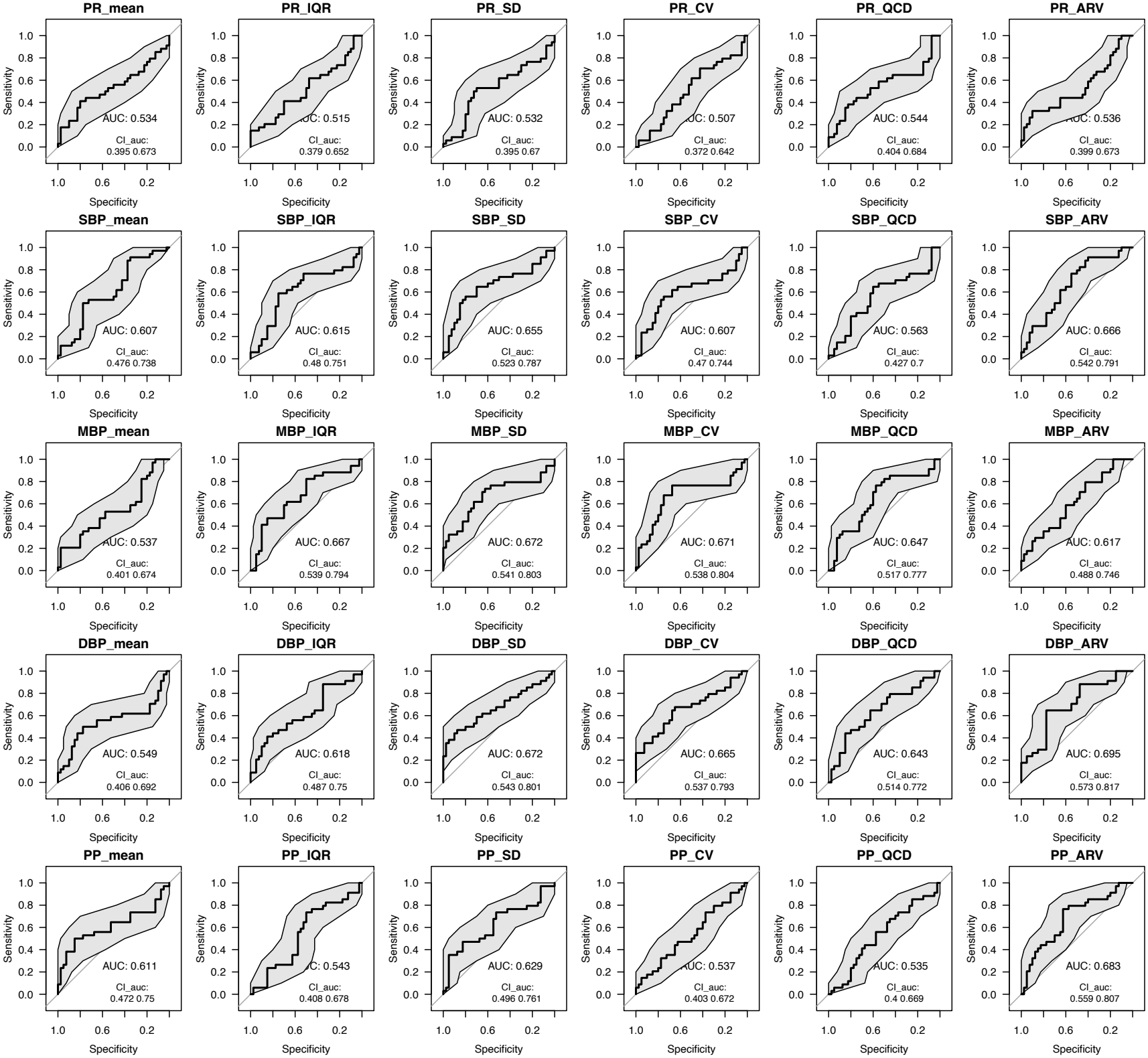
ROC of logistic regression analyses for AKI prediction performance of various blood pressure variability (BPV) parameters with their 95% confidence interval and AUC. ARV: average real variability; AUC: area under the receiver operating characteristic curve; CV: coefficient of variation; DBP: diastolic blood pressure; IQR: interquartile range; MBP: mean blood pressure; PP: pulse pressure; PR: pulse rate; QCD: quartile coefficient of dispersion; SBP: systolic blood pressure; SD: standard deviation.

